# Development and validation of an algorithm to estimate the risk of severe complications of COVID-19 to prioritise vaccination

**DOI:** 10.1101/2021.02.05.21251197

**Authors:** Ron MC Herings, Karin MA Swart, Bernard AM Van der Zeijst, Amber A van der Heijden, Koos van der Velden, Eric G Hiddink, Martijn W Heymans, Reinier AR Herings, Hein PJ van Hout, Joline WJ Beulens, Giel Nijpels, Petra JM Elders

## Abstract

**Objective:** To develop an algorithm (sCOVID) to predict the risk of severe complications of COVID- 19 in a community-dwelling population to optimise vaccination scenarios.

**Design:** Population based cohort study

**Setting:** 264 Dutch general practices contributing to the NL-COVID database

**Participants:** 6074 people aged 0-99 diagnosed with COVID-19

**Main outcome measures:** Severe complications (hospitalisation, institutionalisation, death). The algorithm was developed from a training dataset comprising 70% of the patients and validated in the remaining 30%. Potential predictor variables included age, sex, a chronic co-morbidity score (CCS) based on risk factors for COVID-19 complications as defined by the National Institute of Public Health and the Environment (RIVM), obesity, neighborhood deprivation score (NDS), first or second COVID wave, and confirmation test. Six different population vaccination scenarios were explored: 1) random (*naive*), 2) random for persons above 60 years (*60plus*), 3) oldest patients first in age bands of five years (*oldest first*), 4) target population of the annual influenza vaccination program (*influenza*) and 5) those 25-65 years of age first (*worker*), and 6) risk-based using the prediction algorithm (*sCOVID*). For each vaccination strategy the amount of vaccinations needed to reach a 50% reduction of severe complications was calculated.

**Results:** Severe complications were reported in 243 (4.8%) people with 59 (20.3%) nursing home admissions, 181 (62.2%) hospitalisations and 51 (17.5%) deaths. The algorithm included age, sex, CCS, NDS, wave, and confirmation test with a c statistic of 0.91 (95% CI 0.88-0.94) in the validation set. Applied to different vaccination scenarios, the proportion of people needed to be vaccinated to reach a 50% reduction of severe complications was 67.5%, 50.0%, 26.1%, 16.0%, 10.0%, and 8.4% for the *worker, naive, infuenza, 60plus, oldest first*, and *sCOVID* scenarios respectively.

**Conclusion:** **C**OVID-19 related **s**evere complications will be reduced most efficiently when vaccinations are risk-based, prioritizing the highest risk group using the sCOVID algorithm. The vaccination scenario, prioritising oldest people in age bands of 5 years down to 60 years of age, performed second best. The sCOVID algorithm can readily be applied to identify persons with highest risks from data in the electronic health records of GPs.

**What is already known on this topic?:**

- Severe COVID-19 complications may be reduced when persons at the highest risk will be vaccinated first.
- To identify persons at a high risk for hospitalization or death in the general population, a limited number of prediction algorithms have been developed.
- Most of these algorithms were based on data from the first wave of infections (spring 2020) when widespread testing was not always possible, limiting the usefulness of these algorithms.

**What this study adds:**

- Including data up to January 2021, we developed and validated a prediction algorithm (sCOVID) with a c-statistic of 0.91 (95% CI 0.88-0.94) based on age, sex, chronic comorbidity score, economic status, wave, and a confirmation test to identify patients in the general population that are at risk of severe COVID-19 complication.
- Using the algorithm, a 50% reduction of patients with severe complications could be obtained with a vaccination coverage of only 8%. This vaccination scenario based on this algorithm was superior to other calculated vaccination scenarios.
- The sCOVID algorithm can readily be implemented in the electronic health records of general practitioners.

## Introduction

In the Netherlands, as in many other countries, the SARS-CoV-2 outbreak had severe consequences from March 2020 onwards. The fast spread of the infection and the unexpected severe complications required, in the absence of treatment, hospitalisation for many days in intensive care units (ICU), thereby occupying all available ICU beds in the Dutch hospitals. This urged the Dutch government to install social distancing measures including a lock-down. Although the number of hospitalisations dropped fast in the summer, a sudden increase started in August 2020 leading to a second lockdown on December 15th, 2020 and a curfew on January 23th, 2021. The still limited capacity of available ICU beds, the unpredictable course of the COVID-19 infections, the limited knowledge on how these infections spread among the population, the absence of proper treatments, and the in time and location unsuspected flare-ups of infections, paralysed the Dutch care system and economy.

To prioritise high risk individuals for vaccination or shielding from corona infections, or to start treatment in primary care as soon as possible, accurate identification of patients at risk for severe COVID-19 is of utmost importance. This requires living, accurate risk prediction algorithms, that are easy to apply in General Practice as suggested by Clift et al.^1^. Initially, prediction algorithms for mortality or progression to severe disease were mainly developed for hospitalised patients.^2-4^ In the meantime several prediction algorithms for COVID-19 infected patients in the general population have been developed.^4-6^ Although the performance of these algorithms is fairly good, they have to deal with bias due to country specific policy measures that change in time. This is in part because these studies were conducted based in the first wave of the infections, when testing was scarce and policy measures were still in its infancy.

By now vaccines have become available and vaccination campaigns are ongoing but the shortage of vaccines limits the out roll of these campaigns.^7^ Efforts to prioritise risk groups for vaccination are ongoing, focusing on populations with the highest risk of COVID-19 complications.^8^ The development of our algorithm was aimed to provide predictions for subpopulations at risk for severe COVID-19 infections leading to hospitalisation, institutionalisation or death. The prediction algorithms is based on data of the Dutch NL-COVID database, containing nationwide geo-demographical and medical data. ^9^ Building on this algorithm, that can be updated and adapted regularly, we estimated the effectiveness of six different scenarios for vaccination of high-risk persons in order to prevent severe COVID-19 complications.

## Methods

### Design

This cohort study was performed by using data from an extensive and representative general practice population database in the Netherlands.

### Data sources

Data were obtained from GP practices who reported information of the diagnoses and comorbidities of patients suffering from COVID-19 in the NL-COVID database. This database was set up in April 2020 as a collaborative initiative of general practitioners, public health specialists, virologists, epidemiologists, data scientists, data specialists, privacy specialists, and ICT companies providing Electronic Health Records (EHR). Together the ICT companies cover about 95% of all GP practices in the Netherlands. GPs were asked to complete a brief questionnaire protocol for patients suffering from COVID-19 in their ICT systems. From a total of 264 practices (∼5% of all Dutch GP practices), both questionnaire data and EHR records with information regarding selected comorbidities were included in this study. Data until January 21th, 2021 were used.

The selected comorbidities (Supplementary Table 1) were those indicated by the National Institute for Public Health and the Environment (RIVM) to be relevant for the prognosis of severe outcomes of COVID-19 infections.^10^ The following information was collected on a daily basis: a diagnosis of COVID-19 and whether the diagnosis was confirmed with a PCR test, the severity of the infection defined as treated at home, treated in a hospital or special care institution or death from COVID-19. Updates of the patient’s status was recorded using the same form. For this paper we used the last status report. In addition, age, gender, body mass index (BMI), and postal code were collected from the electronic registries of the GP. The neighborhood deprivation score (NDS) was based on the quartile distribution of relative wealth of the neighborhood as derived by Statistics Netherlands.^11^ There were no missing data in the NL-COVID database: questionnaire data were complete and the registration of comorbidities in the EHR was considered to be complete as well.

### Participants

A cohort study was performed among patients registered in the NL-COVID database suffering from COVID-19 symptoms certified by their GP.

### Primary outcome

The primary outcome was the occurrence of severe complicated COVID-19 disease defined as hospitalisation, institutionalization or death.

### Predictors

Predictors included age and sex, the NDS, body mass index (BMI ≥ 30 kg/m^2^), the period of registration (before or after August 2020) as first of the second wave, whether the diagnosis was confirmed with a PCR test or CT-Scan, and a chronic comorbidity score (CCS). The CCS was based on the chronic diseases identified as predictors for complications of COVID-19 infection by the National Institute for Public Health and the Environment.^10^ The comorbidities were mapped to the ICPC coding system used in Dutch GP practices and subsequently grouped into nine disease clusters (Appendix A). A patient was scored in each of these respective disease clusters and assigned a point per cluster. For example, a patient suffering from epilepsy and diabetes scored a point in the category neurological diseases and a point for diabetes yielding a CCS of 2.

### Risk mitigation scenarios

The prediction models yield a probability that a COVID-19 patient develops a severe complication. In a single normalized Dutch GP practice (N= 2090 patients) the summarised sCOVID predicted probabilities is 85. It is assumed that if all patients would be infected with COVID-19, an expected 85 patients would develop severe COVID-19 complications. We further assumed that this probability can be reset to (almost) zero by vaccination, or by shielding patients from contact with others. By vaccination or shielding of the 10 highest-ranked patients, ranging from a probability 0.64 to 0.45, 85 minus 5 = 80 patients were expected to develop severe COVID-19 complications, a decrease of 100 * (1- 80/85) of 5.9%. For each GP practice, the predicted number of patients developing severe COVID-19 complications is estimated as the summarized sCOVID probabilities as Base (B). Depending on the vaccination coverage and the policy who to vaccinate (scenario), the number of patients developing severe complications can be estimated for different vaccination scenarios.

The impact of the vaccination strategy can be followed in time by division of the summarized probabilities Pt divided by B as 100% times Pt/B yielding the percentage expected decrease in severe complications at a given percentage of the population vaccinated. The vaccination coverage needed for a 50% decrease of hospitalization was defined as VC50 as a measure of the efficiency of a particular hypothetical vaccination or shielding scenario. We explored and compared six different hypothetical vaccination scenarios. A first scenario was defined as a *naive* scenario, a scenario in the absence of any policy, i.e. inhabitants are randomly vaccinated. A second scenario was defined as a *plus60* scenario where all inhabitants, 60 years of age or older are randomly vaccinated, followed by random vaccination of those under 60 years of age (also random). A third scenario (*oldest first*) prioritised vaccination from the oldest down from 100 to 60 years of age in age bands of 5 years. Within the respective age bands, allocation is random. A fourth scenario was defined as the *influenza* scenario. Here, patients with an indication for flu vaccination are prioritized for vaccination. A fifth scenario (*worker*) prioritized random vaccination of inhabitants 25-65 years of age. The six and the last scenario was based on the sCOVID risk ranking algorithm, the *sCOVID* scenario. Here, we start vaccination based on the absolute risk ranking, the patient with the highest risk first, followed by the second patient in line etc etc.

### Statistical analyses

LASSO regression analysis was used to select predictors in the model and to estimate and shrink regression coefficients. Ten-fold cross validation was used to estimate the optimal shrinkage factor (λ) used in the LASSO regression, such that the sum of the squared residuals was minimised. Age was included as quadratic function. The final regression formula allowed calculation of predicted probabilities for each registered patient at their GP. We randomly allocated 70% of the patients in a training dataset to develop the model. The other 30% of the patients was allocated into a validation dataset. We assessed the model performance in terms of discrimination and calibration in the validation set. Discrimination was assessed using the c-statistic. The c-statistic indicates the extent to which the model can distinguish between a patient with and without the outcome and varies between 0.5 and 1. Calibration was assessed using calibration plots showing the predicted risk against the observed frequency of the study population’s outcome using ten risk groups. Goodness of fit was assessed with the Brier score to quantify the difference between the observed and fitted probability ranging for 0-1 with a score of 0 representing the best model.^12^ R version 4.0.2., GLMNET package (4.0-2), version was used for statistical analyses and constructing figures. We adhered to the Transparent Reporting of a multivariable prediction model for Individual Prognosis Or Diagnosis (TRIPOD) statement.^13^

### Patient and public involvement

Patients were not involved in the design and conduct of the study. General practitioners were consulted to reflect on their ideas about different vaccination scenarios and their practicality.

## Results

### Overall study population

A total of 264 GP practices (∼5% of all Dutch GP practices) reported 6074 patients with a diagnosis of COVID-19 in the period 10th April 2020- until 21st January 2021. Severe complications were reported for 291 (4.7%) patients of whom 59 (20.3%) was treated in a nursing home, 181 (62.2%) were hospitalized and 51 (17.5%) patients died. Training and test model included 4251 and 1823 persons, respectively.

### Baseline characteristics

The characteristics of COVID-19 patients recorded in the first and second time period differed in age, baseline risk, frequency of testing and region. The percentage of people developing severe complications dropped from 8.5% in the first period in the Spring 2020 to 2.5% in the second time period in the Autumn 2020 which was reflected in institutionalisation, hospitalisation, and death. In the first wave, infected patients were from older age groups, whereas relatively more adolescents, 12- 19 years of age, were reported in the second wave. The proportion of patients recorded with a positive COVID-19 test increased from 63% in the first wave to 95% in the second wave. The general characteristics are presented in Table 1. Most of the patients with severe complications suffered from cardiovascular conditions (64.6%) and other chronic conditions such as diabetes, neurological diseases (i.e., dementia, Parkinson’s diseases) and lung disease. Almost 80% of patients with severe complications suffered from at least one chronic disease. More than 62% had multiple chronic conditions. The characteristics of the training and validation set is shown in table 2.

**Table 1:** Characteristics of the cohort of COVID-19 patients.

|  | Characteristic | Complicated (%)<br>(N=291) | Home (%)<br>(N=5783) |
| --- | --- | --- | --- |
| Outcome | Nursing home | 59 (20.3%) | - |
|  | Hospitalisation | 181 (62.2%) | - |
|  | Deceased | 51 (17.5%) | - |
| Age group | <40 yrs | 17 (5.8%) | 2186 (37.8%) |
|  | 40-49 yrs | 16 (5.5%) | 985 (17.0%) |
|  | 50-59 yrs | 34 (11.7%) | 1204 (20.8%) |
|  | 60-69 yrs | 43 (14.8%) | 758(13.1%) |
|  | 70-79 yrs | 66 (22.7%) | 436 (7.5%) |
|  | 80-89 yrs | 79 (27.1%) | 178 (3.1%) |
|  | 90+ yrs | 36 (12.4%) | 36 (0.6%) |
| Sex | Woman | 144 (39.5%) | 3352 (58.0%) |
|  | Man | 147 (50.5.1%) | 2431 (42.0%) |
| Timing | Period before August | 197 (67.7%) | 2123 (36.7%) |
|  | Period starting in August | 94 (32.3%) | 3660 (63.3%) |
| Province | South Holland | 92 (31.6%) | 2102(36.3%) |
|  | North Brabant | 82 (28.2%) | 2495 (43.1%) |
|  | North Holland | 67 (23%) | 821 (14.2%) |
|  | Limburg | 33 (11.3%) | 227 (3.9%) |
|  | Other | 17(5.8%) | 138 (2.4%) |
| Tested | Negative or not | 49 (16.8%) | 1025 (17.7%) |
|  | Positive | 242 (83.2%) | 4758 (82.3%) |
| NDS | Middle | 85 (29.2%) | 2241 (38.8%) |
|  | Low | 150 (51.5%) | 1900 (32.9%) |
|  | High | 56 (19.2%) | 1642 (28.4%) |
| Obesity | BMI ≥ 30 kg/m2) | 50 (17.2%) | 611 (10.6%) |
| Comorbidity | Cancer | 84 (28.9%) | 507 (8.8%) |
|  | Cardiovascular disease | 188 (64.6%) | 1405 (24.3%) |
|  | Diabetes | 64 (22.0%) | 440 (7.6%) |
|  | Heart valve disease | 21 (7.2%) | 87 (1.5%) |
|  | Immunosuppresants | 41 (14.1%) | 421 (7.3%) |
|  | Kidney disease | 67 (23.0%) | 280 (4.8%) |
|  | Liver disease | 7 (2.4%) | 89 (1.5%) |
|  | Lung disease | 64 (22.0%) | 864 (14.9%) |
|  | Neurological disease | 42 (14.8%) | 187 (3.2%) |
| Indication | Influenza vaccination | 225 (77.3%) | 2392 (42.3%) |
| CCS | 0 | 60 (20.6%) | 3250 (56.2%) |
|  | 1 | 49 (16.8%) | 1430 (24.7%) |
|  | 2 | 74 (25.4%) | 655 (11.3%) |
|  | 3 | 65 (22.3%) | 297 (5.1%) |
|  | 4 | 31(10.7%) | 113(2.0%) |
|  | 5+ | 12 (4.1%) | 38 (0.7%) |

CCS= chronic comorbidity score, NDS = neighborhood deprivation score

**Table 2:** Characteristics of training and test cohort of COVID-19 patients.

| Predictors | Sampling | Training (70% Random) |  | Test (30% Random) |  |
| --- | --- | --- | --- | --- | --- |
|  |  | Complicated | Home | Complicated | Home |
|  |  | (N=197) | (N=4054) | (N=94) | (N=1729) |
| Age group | <40 yrs | 15 (7.6%) | 1514 (37.3%) | 2 (2.1%) | 672 (38.9%) |
|  | 40-49 | 14 (7.1%) | 682 (16.8%) | 2 (2.1%) | 303 (17.5%) |
|  | 50-59 | 21 (10.7%) | 850 (21.0%) | 13 (13.8%) | 354 (20.5%) |
|  | 60-69 | 33 (16.8%) | 539 (13.3%) | 10 (10.6%) | 219 (12.7%) |
|  | 70-79 | 42 (21.3%) | 321 (7.9%) | 24 (25.5%) | 115 (6.7%) |
|  | 80-89 | 49 (24.9%) | 122 (3.0%) | 30 (31.9%) | 56 (3.2%) |
|  | 90+ | 23 (11.7%) | 26 (0.6%) | 13 (13.8%) | 10 (0.6%) |
| Sex | Man | 87 (44.2%) | 1711 (42.2%) | 57 (60.6%) | 720 (41.6%) |
|  | Woman | 110 (55.8%) | 2343 (57.8%) | 37 (39.4%) | 1009 (58.4%) |
| Timing | Before August 2020 | 59 (29.9%) | 2587 (63.8%) | 35 (37.2%) | 1073 (62.1%) |
|  | After august 2020 | 138 (70.1%) | 1467 (36.2%) | 59 (62.8%) | 656 (37.9%) |
| Tested | Negative or not | 32 (16.2%) | 698 (17.2%) | 17 (18.1%) | 327 (18.9%) |
|  | Positive | 165 (83.8%) | 3356 (82.8%) | 77 (81.9%) | 1402 (81.1%) |
| NDS | Middle | 58 (29.4%) | 1568 (38.7%) | 27 (28.7%) | 673 (38.9%) |
|  | Low | 102 (51.8%) | 1346 (33.2%) | 48 (51.1%) | 554 (32.0%) |
|  | High | 37 (18.8%) | 1140 (28.1%) | 19 (20.2%) | 502 (29.0%) |
| Overweight | BMI<30 | 164 (83.2%) | 3621 (89.3%) | 77 (81.9%) | 1551 (89.7%) |
|  | BMI ≥30 | 33 (16.8%) | 433 (10.7%) | 17 (18.1%) | 178 (10.3%) |
| CCS | 0 | 42 (21.3%) | 2273 (56.1%) | 18 (19.1%) | 977 (56.5%) |
|  | 1 | 40 (20.3%) | 1010 (24.9%) | 9 (9.6%) | 420 (24.3%) |
|  | 2 | 52 (26.4%) | 471 (11.6%) | 22 (23.4%) | 184 (10.6%) |
|  | 3 | 39 (19.8%) | 198 (4.9%) | 26 (27.7%) | 99 (5.7%) |
|  | 4 | 19 (9.6%) | 73 (1.8%) | 12 (12.8%) | 40 (2.3%) |
|  | 5+ | 5 (2.5%) | 29 (0.7%) | 7 (7.4%) | 9 (0.5%) |

CCS= chronic comorbidity score, NDS = neighborhood deprivation score

### Predictor variables

The predictor variables in the final COVID-19 models included age, sex, positive test result, period (first or second wave), NDS, obesity and the CCS (Table 3). The strongest predictors included age, NDS, the time period, a positive PCR test and male sex. Obesity was eliminated by the LASSO predictor selection. The final model showed a very good calibration and fit. Figure 1 illustrates the ROC curve from the validation set with a c-index of 0.91 (95%CI: 0.88-0.94). The model yielded a good calibration (Brier score= 0.034) (Figure 2).

**Table 3:** Selected predictors of LASSO regression coefficients.

| Predictors | LASSO Coefficients |
| --- | --- |
| Intercept | -5.1765361941 |
| Age (squared) | 0.0005142466 |
| Male | 0.1133382856 |
| After July 2020 | -1.3845037172 |
| Positive COVID-19 test | 0.6423757757 |
| Obese (> 30 kg/m2) | . |
| Low NDS | 0.4400574635 |
| High NDS | . |
| CCS | 0.1186100356 |

CCS= chronic comorbidity score, NDS= neighborhood deprivation score

**Figure 1.**
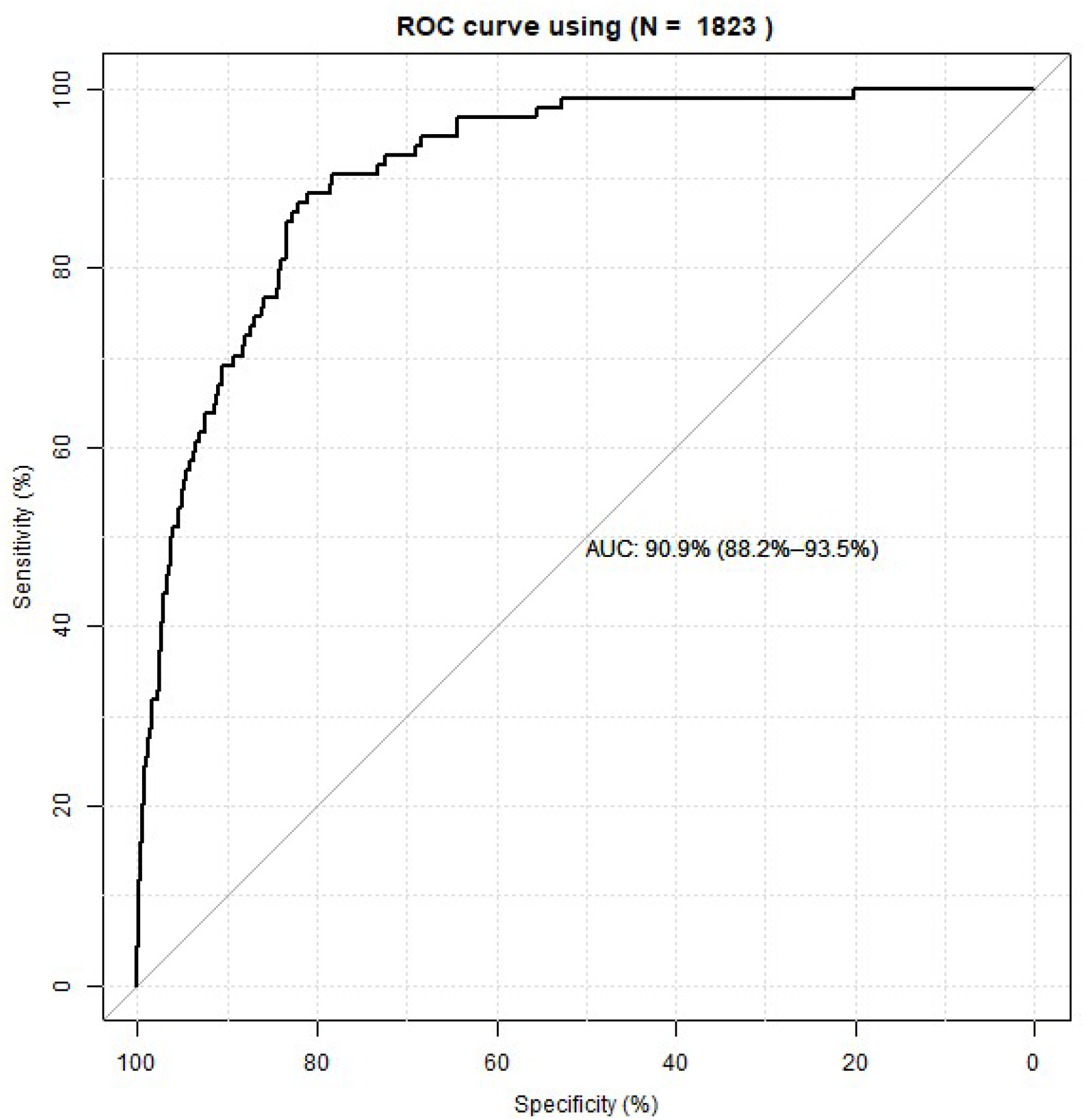
ROC Curve LASSO Regression. Receiver operation characteristic curve of sCOVID based on the validation samples. The c- index was 0.91.

**Figure 2.**
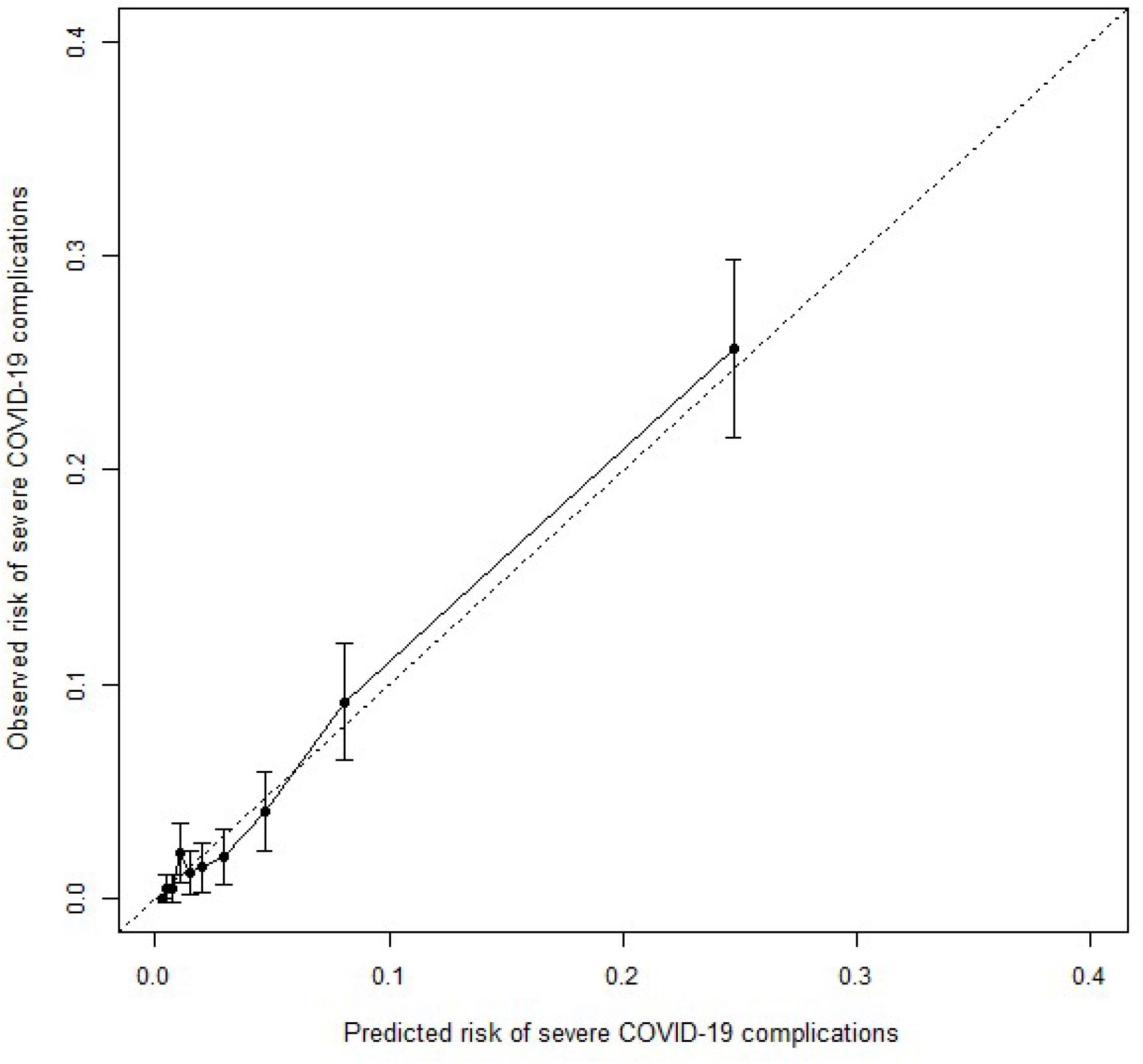
Calibration Plot LASSO Regression. Plot of severe COVID-19 complications predicted by the sCOVID algorithm versus the observed complications. The Brier score is 0.034.

**Figure 3:**
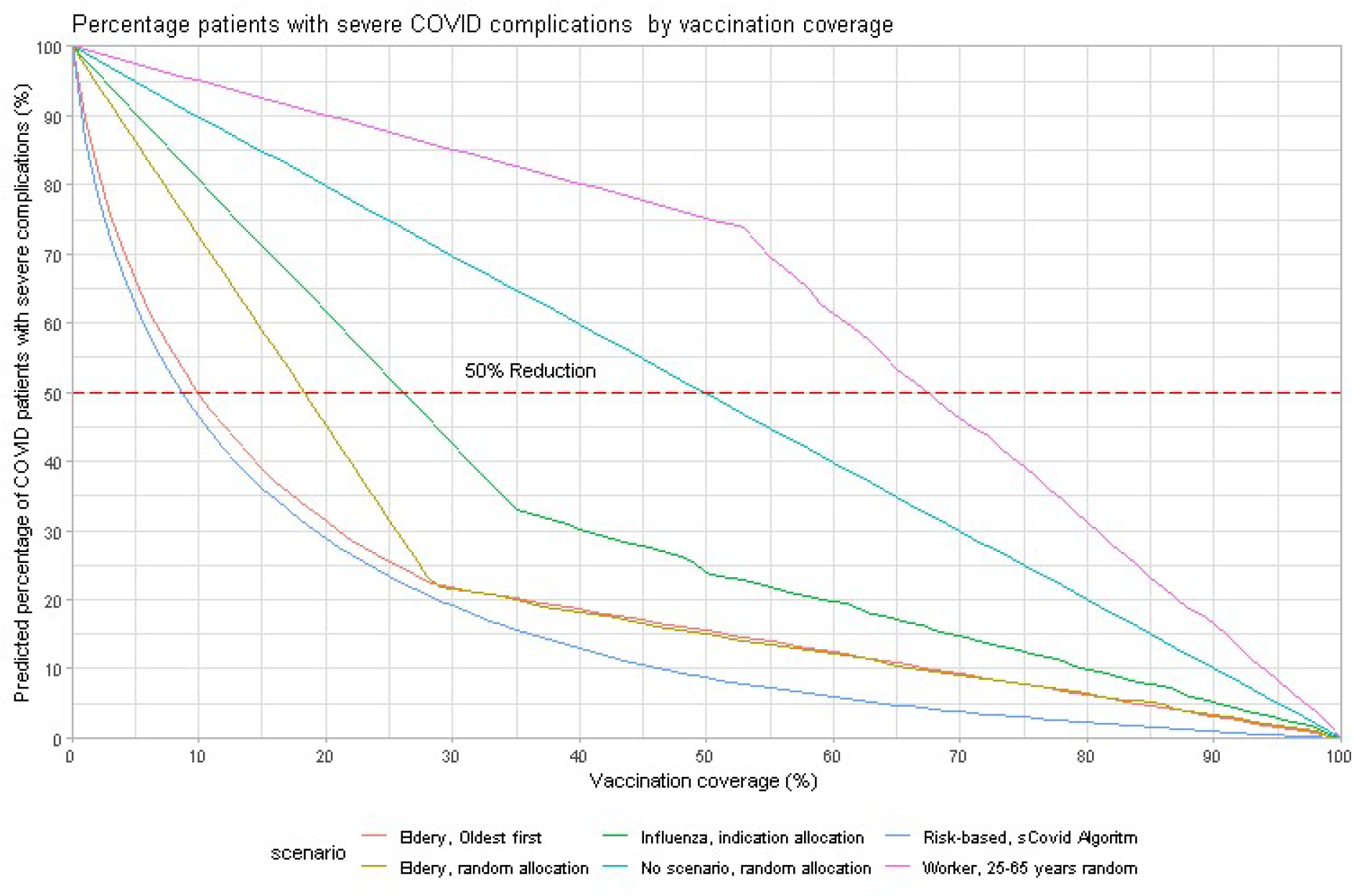
Results of different risk stratification scenarios. Effect of vaccination on the reduction of severe COVID-19 complications in different scenarios. Vaccination coverages needed for a 50% reduction of the burden of disease can be estimated at the intersection of the graphs with the 50% Reduction line.

### Risk prediction in practice

#### Examples of individual risk ranking

The risk of developing severe complications for a 60-year-old man, with a positive PCR test, living in a neighborhood with a low NDS, who suffers from diabetes, hypertension and kidney failure can be estimated. His comorbidities comprise three different classes (Appendix A). Summarizing the coefficients (Cf) from the column LASSO regression of Table 3, the equation yields as total score of Cf(intercept) + 60*Cf (age times age) + Cf(man) + Cf (after July 2020) + Cf (positive COVID-19 test) + Cf (low NDS) + 3*Cf (CCS) = −3.456. His risk to develop severe complications is subsequently calculated as 100*(exp (−3.456) / (1 + exp (−3.456) = 4.1%. The risk equals that of a 73-year-old woman without any chronic condition living in a neighborhood with a high socioeconomic status.

### Practice risk ranking

The results of six prioritising scenario analyses were obtained by deploying the different algorithms to 300 randomly selected, fully anonymised GP practices, including data from 1.2 million inhabitants. The results for the six scenarios are plotted in Figure 5 and summarized in Table 4. A reduction of 50% of the patients with severe complications was observed already with a vaccination coverage of 8% if all high-risk persons according to the sCOVID algorithm are vaccinated first. This scenario was superior to all other scenarios with vaccination scheme in which the oldest are consecutively vaccinated in age bands of five years being second best. The worst scenario were the *worker* scenario prioritising patients 25-65 years of age, followed by the naïve scenario where patients are randomly vaccinated.

**Table 4:** Vaccination coverage needed for 50% risk reduction by selected scenarios.

| Scenario | Strategy | Vaccination coverage for 50% Risk reduction | Needed vaccinations in an normalized sized GP practice* (Netherlands ) |
| --- | --- | --- | --- |
| sCOVID based | Highest risk first, based on ranking from highest to lowest risk | 8.4% | 175 (1.2 million) |
| Oldest first | Stepwise in 5 years group from 100 to 65, random in ageband | 10.0% | 210 (1.7 million) |
| 60 plus | 60 plus first, random | 18.1% | 380 (3.1 million) |
| Influenza | Those with an indication for influenza vaccination first, random | 26.1% | 545 (4.4 million) |
| Naive | None, random, everybody | 50.0% | 1050 (8.5 million) |
| Worker | Prioritize inhabitants 25-65 years of age | 67.5% | 1414 (11.5 million) |

\*Normalized GP practice = 2095 patients (www.lhv.nl)

## Discussion

Using data from the NL-COVID database an algorithm was developed to predict the probability of patients developing severe complications once infected with COVID-19 using EHR records from general practices. This sCOVID algorithm, which can be deployed in all Dutch GP practices, showed a very good performance in terms of discrimination (c-index: 0.91) and calibration and can be used to rank the most susceptible patients for prioritisation of vaccinations. Our vaccination scenarios showed that ranking and vaccinating patients based on their complication risk (sCOVID scenario) would be the most efficient vaccination scenario to reduce hospitalisation and deaths. The second-best scenario was to vaccinate the oldest people first in consecutive order. With shortage of vaccines, the most vulnerable patients and not the oldest patients are prioritized.

### Comparison with other studies

The sCOVID risk models yields a high discrimination rate (c-statistic = 0.91). Calibration plots show a good fit in all risk categories, although the lowest risks were most challenging to estimate due, to the limited numbers of patients developing severe complications. These results are similar to those of other prediction algorithms. Two earlier studies developed prediction algorithms for hospitalisation and/or death due to COVID-19 infection show similar prognostic performance.^4 5^

The major predictors, selected by the LASSO procedures, were higher age, male gender, the number of chronic comorbidities but also a positive test result and neighborhood deprivation status. These selected predictors resemble the predictors reported in earlier studies by Clift et al., Jehi et al and Williamson et al.^4 5 6^ The most obvious differences were the summary score of comorbidities (CCS) compared to separate conditions and inclusion of symptoms and laboratory measures for the study by Jehi et al. Most predictors found in this study relate to poor health and a complex of comorbidities. More than 60% of the patients with severe complications suffered from more than one chronic condition against less than 20% of the those without comorbidities. Therefore, we preferred to include a chronic disease summary score to come to a more comprehensive and practical algorithm. Moreover, from a clinical perspective, our sample size was relatively small and would exclude rare but clinically relevant outcomes.

### Complexity of modelling

Estimating the risk of severe COVID-19 complications is permanently subject to changing policy measures and interventions to shield high-risk people by vaccinations.^7,8^ The time biases caused by these measures and interventions are complex and difficult to unravel. First analyses confirmed suggestions from Clift et al. that these time biases are indeed present,^4^ showing an age and sex-adjusted 3-5 times lower complication rate compared to the patients in the first wave. Estimates, needed to predict hospitalisation and or death, therefore need permanent recalibration of the prediction algorithms. Such recalibration is necessary to monitor the effect of policy intervention on managing care capacity. The infrastructure of the NL-COVID database permits the recalibration on a regional and daily basis.

### Strengths and limitations

The NL-COVID database also has limitations and strengths. First, we have substantial underreporting of positive cases since our 264 registration practices consisting of about 5% of all GP practices only reported 0.7% of the registered cases. This is explained by several factors: first, practices enrolled into the program over time and some practices only joined the program and the end of 2021. Second, COVID-19 testing was done by the regional health authorities (GGD) whereas the administrations of the GGD were not linked with the GP administration. Therefore our registration relies on whether the patient contacted the GP and whether the GP registered the patient. This makes it likely that we have a selection bias towards the more severe disease manifestations of the COVID-19 infection. Also our prediction partly relied on the judgement of the GP whether a patient was COVID 19 positive (in case of lacking test results). It should therefore be stressed that absolute estimates risk estimates to develop severe complications should be interpreted with care only by health care professionals for prioritizing strategies. A weakness of the sCOVID scenario is that we did not perform an external validation. The large number of GP practices that came from all over the country and the good testing characteristics of the validation set, makes it likely that the accuracy of the scenarios is adequate. For the comparison of the different scenarios this has no importance since they were compared in the same sample.

A first strength was the coverage and representativity of the practices most strongly confronted with the pandemic. The first wave of COVID-19 hit hard in the southern part of the country and most participating practices were situated here. Second, we used training and validations samples to estimate the accuracy of the algorithms. Third, this study is the first to demonstrate the potential impact and efficiency of more and less targeted vaccinations scenarios. Fourth, the prediction algorithm can be adapted, updated and validated on a daily basis and learn from new insights and policy measures.

### Practical implications

Our study showed that within the framework of privacy regulations, COVID-19 infections and consequences can be monitored fast, efficiently and safe on a very detailed local levels and on a day-to-day basis using country wide data from currently available IT systems in GP practice. The costs of such a database are relatively low. Insight can be generated that help GPs and involved regional and local health authorities to shield patients from infection and to reduce hospitalisation and death very efficiently in a selected group of persons with the highest risks. Moreover, such database may demonstrate and underpin the effectiveness and efficiency of policy measures, to plan and manage care facilities. Secondly, the prediction accuracy could be improved with flexible access to the complete GP patient dossier and linkage to hospital admission under strict compliance with the GDPR to adapt and improve the algorithms if new insights become available. The vaccination scenarios show that in case of remaining shortage of vaccines, vaccination base on the sCOVID scenario performs best with a consecutive age based scenario as second best. Hybrid scenarios that do not follow the risk of COVID-19 complications have worse performances for example the influence scenario in which a combination of age and influenza risk is combined. Currently, vaccination in the Netherlands is performed from a practical perspective based on factors not only related to COVID risk complications. This makes it likely that more efficient scenarios are thinkable. In conclusion, the sCOVID algorithm has been developed to predict which patients are at high risk to develop severe complications due to COVID-19 and showed a good model performance. In remaining shortage of vaccines prioritizing vaccination of patients based on sCOVID risk complications is the most efficient way to reduce hospitalisations, institutionalisations and death.

## Supporting information

TRIPOD checklist

## Data Availability

The data will be available on reasonable request.

## Acknowledgement

The authors like to thank the unconditional support of among which Guus Vaassen and Johan Ruiter (Medworq), Marjoleine van der Zwan, Piet-Hein Knoop, Arjan den Ouden (Pharmapartners), Mark van Vliet, Chris Tromp (Health Base foundation), Eric Grosveld, Meefa Hogenes (Expertdoc), and Frank Carlebur (ZonH), Ernst de Graag, Michiel Meulendijk (STIZON), Theo Peters (CGM) and more than 450 Dutch general practitioners that contribute to the COVID-Database in time, cash or kind to fight COVID-19.

## Additional information

### Contributorship

RMCH, EGH were involved in the development of the database. RMCH, KMAS, BAMvdZ, AAvdH, KvdV, HvH, JWJB, GN, and PMJE contributed to the development of the research question and study design. RMCH and RARH conducted the statistical analyses. MWH was involved in advanced statistical aspects. All authors contributed to the interpretation of the results. RMCH wrote the first draft of the manuscript. All authors contributed to the critical revision of the manuscript for important intellectual content and approved the final version of the manuscript. RMCH is the guarantor.

### Ethical approval

Patients and GPs were asked to consent with data sharing regarding sending data to the NL-COVID database and extract information on the 4-digit postal code level in an anonymized format for public decision making. The procedure was approved and tested for compliance with the general data protection regulation by the privacy and compliance committee of ‘Stichting Informatie voorziening voor Zorg en Onderzoek’ (STIZON).

### Transparency declaration

The lead author affirms that the manuscript is an honest, accurate, and transparent account of the study being reported; that no important aspects of the study have been omitted; and that any discrepancies from the study as originally planned (and, if relevant, registered) have been explained.

### Data sharing

The data will be available on reasonable request.

