## Appendix A for "Development and validation of an algorithm to estimate the risk of severe complications of COVID-19 to prioritise vaccination"

### Appendix A: Characteristics and risk factors selected for the development of the sCOVID prediction model

| Type of data | Characteristic | ICPC/ Diagnostic code/ ATC |
|---|---|---|
| **Demographic** | Age in years (continuous) | |
| | Sex | |
| | NDS (Area postal level) | |
| | Postal code (4 – digits) | |
| **Lifestyle** | | |
| *Obesity* | Obesity (BMI ≥ 30 kg/m2) | T82 |
| | BMI ≥ 30 kg/m2 in the last 2 years | 1272 |
| **COVID-19 (Information extracted from questionnaire)** | | |
| *Current Status* | Clinical suspicion (not tested) | |
| | COVID-19 test positive (any) | |
| *Severity* | Hospitalisation | |
| | Institutionalisation | |
| | Deceased | |
| *Time period* | First wave (March - August 2020) | |
| | Second wave (September-December 2020) | |
| **Chronic Diseases (CMS: score 0- 9)*** | | |
| *Cardiovascular* | Ischaemic heart disease with angina | K74 |
| | Unstable angina | K74.01 |
| | Stable angina | K74.02 |
| | Acute myocardial infarction | K75 |
| | Ischaemic heart disease without angina | K76 |
| | Coronary sclerosis | K76.01 |
| | Old heart attack | K76.02 |
| | Heart failure | K77 |
| | Acute heart failure | K77.01 |
| | Chronic heaft failure | K77.02 |
| | Atrial fibrillation/flutter | K78 |
| | Transient cerebral ischaemia | K89 |
| | Stroke/cerebrovascular accident | K90 |
| | Subarachnoid hemorrhage | K90.01 |
| | Intracerebral haemorrhage | K90.02 |
| | Cerebral Infarction | K90.03 |
| | Cerebrovascular disease | K91 |
| | Hypertension uncomplicated | K86 |
| | Hypertension complicated | K87 |
| | Paroxysmal tachycardia | K79 |
| | Supraventricular tachycardia | K79.01 |

| | | |
|---|---|---|
| | Ventricular tachycardia | K79.02 |
| | Congenital anomaly cardiovascular | K73 |
| | Atrial septal defect | K73.01 |
| | Ventricular septal defect | K73.02 |
| | Heart disease other | K84 |
| | Wolff-Parkinson-White (WPW) syndrome | K84.01 |
| | Atrioventricular block | K84.02 |
| | Cardiomyopathy | K84.03 |
| | Long QT syndrome | K84.07 |
| | Atherosclerosis/peripheral vascular disease | K92 |
| | Intermittent claudication | K92.01 |
| | Raynaud syndrome | K92.02 |
| | Buerger's disease | K92.03 |
| | Cardiac arrhythmia NOS | K80 |
| | Supraventricular extrasystoles | K80.01 |
| | Ventricular extrasystoles | K80.02 |
| | Sick sinus syndrome | K80.03 |
| | Pulmonary heart disease | K82 |
| *Diabetes* | Diabetes non-insulin dependent | T90 |
| | Diabetes mellitus type 1 | T90.01 |
| | Diabetes mellitus type 2 | T90.02 |
| *Neurological* | Dementia | P70 |
| | Parkinsonism | N87 |
| | Parkinson's disease | N87.01 |
| | Epilepsy | N88 |
| | Multiple sclerosis | N86 |
| | Neurological disease other | N99 |
| | ALS | N99.01 |
| | Myasthenia | N99.02 |
| | Other neuron muscle diseases | N99.03 |
| *Heart valve* | Rheumatic fever/heart disease | K71 |
| | Rheumatic and heart disease | K71.02 |
| | Heart valve disease NOS | K83 |
| | Stenosis of aorta | K83.01 |
| | Mitral insufficiency | K83.02 |
| *Liver* | Liver disease NOS | D97 |
| | Cirrhosis | D97.04 |
| | Liver steatosis (< 5 years) | D97.05 |
| | Viral hepatitis | D72 |
| | Viral hepatitis A (< 6 month) | D72.01 |
| | Viral hepatitis B (< 6 month) | D72.02 |
| | Viral hepatitis C (< 6 month) | D72.03 |
| | Carrier Hepatitis B | D72.04 |
| | Carrier Hepatitis C | D72.05 |

| | | |
|---|---|---|
| *Lung* | Chronic obstructive pulmonary disease | R95 |
| | Astma | R96 |
| | Allergic astma | R96.02 |
| | Chronic bronchitis | R91 |
| | Chronic bronchitis | R91.01 |
| | Bronchiectasis | R91.02 |
| | Pulmonary embolism | K93 |
| | Tuberculosis | R70 |
| | Pleurisy/pleural effusion | R82 |
| | Congenital anomaly respiratory | R89 |
| | Respiratory disease other | R99 |
| | Pneumoconiosis | R99.06 |
| | Cystic fibrosis | T99.10 |
| | GOLD classification (<24 month) | 2209 |
| | Nr of exacerbations (<12 month) | 3549 |
| | Prednisolone (<24 month) | H02AB06 |
| | Prednisone (<24 month) | H02AB07 |
| *Cancer* | Malignant neoplasm breast female | X76 |
| | Malignant Adenocarcinoma | X76.01 |
| | Malignant neoplasm male genital other | Y78 |
| | Malignant carcinoma penis | Y78.01 |
| | Malignant carcinoma testis | Y78.02 |
| | Malignant carcinoma breast | Y78.03 |
| | Malignant neoplasm nervous system | N74 |
| | Malignant neoplasm of bladder | U76 |
| | Malignant neoplasm thyroid | T71 |
| | Malignant neoplasm pancreas | D76 |
| | Malignant neoplasm stomach | D74 |
| | Malignant neoplasm related to pregnancy | W72 |
| | Malignant neoplasm urinary tract other | U77 |
| | Malignant neoplasm of kidney | U75 |
| | Malignant neoplasm colon/rectum | D75 |
| | Malignancy NOS | A79 |
| | Malignant neoplasm cervix | X75 |
| | Malignant neoplasm bronchus/lung | R84 |
| | Malignant neoplasm blood other | B74 |
| | Multiple myeloma | B74.01 |
| | Malignant neoplasm genital female other | X77 |
| | Endometrial carcinoma | X77.01 |
| | Malignant ovary carcinoma | X77.02 |
| | Malignant neoplasm of skin | S77 |
| | Basal cell carcinoma | S77.01 |
| | Spinocellular carcinoma | S77.02 |
| | Malignant melanoma | S77.03 |

| | | |
|---|---|---|
| | Kaposi's sarcoma | S77.04 |
| | Malignant neoplasm respiratory other | R85 |
| | Benign neoplasm respiratory | R86 |
| | Hodgkin Lymphoma | B72 |
| | Hodgkin Lymphoma | B72.01 |
| | Non-hodgkin Lymphoma | B72.02 |
| | Leukaemia | B73 |
| *Kidney* | Congenital anomaly urinary tract | U85 |
| | Polycystic kidney disease | U85.01 |
| | Glomerulonephritis/nephrosis | U88 |
| | Urinary disease other | U99 |
| | Chronic kidney disease | U99.01 |
| | Renal hypoplasia | U99.02 |
| | Hydronephrosis | U99.03 |
| | Creatin clearance (CKD-EPI) <30 (< 12 months) | 3583 |
| | Creatin clearance (MDRD) <30 (< 12 months) or | 1919 |
| | Creatin clearance (COCKCROFT) <30 (< 12 months) or | 1918 |
| | Creatin clearance (< 12 months) | 524 |
| *Immune system* | HIV-infection/AIDS | B90 |
| | Seropositive without symptoms | B90.01 |
| | HIV | B90.02 |
| | Immunodeficiencies | T99.01 |
| | Prednison/prednisolon (< 6 months) | H02A* |
| | Oncolytics (< 6 months) | L01* |
| | Immunosuppressants (< 6 months) | L04A* |
| | TNF-Alpha (< 6 months) | L04AB01 |
| | Interleukine inhibititors (< 6 months) | L04AC |
| **Indication for influenza vaccination (score 0-1)** | | |
| | Diabetes | |
| | Cardiovascular disease | |
| | Lung disease | |
| | Kidney disease | |
| | Immunosuppressive disease/immunosuppressants | |
| | Heart valve disease | |
| | Cirrhosis | D97.04 |
| | Liver disease NOS | D97 |
| | Congenital anomaly NOS/multiple | A90 |
| | Down syndrome | A90.01 |
| | Ruptured spleen traumatic | B76 |
| | Hereditary hemolytic anemia | B78 |
| | Sikkelcell anemia | B78.02 |
| | Congenital anomaly musculoskeletal | L82 |
| | Acquired deformity of the spine | L85 |

| | | |
|---|---|---|
| | Scoliosis | L85.01 |
| | Endocrine/metabolic/nutritional disease other | T99 |
| | Cushing syndrome | T99.08 |
| | Addison syndrome | T99.09 |
| | Adrenal insufficiency | T99.12 |
| | Adreno-genital syndrome | T99.13 |

**A patient with dementia, epilepsy and diabetes generates a score of 3*
